# Domain Adaptation Strategies for Transformer-Based Disease Prediction Using Electronic Health Records

**DOI:** 10.1101/2025.06.02.25328621

**Authors:** Johanna Driever, Manuel Lentzen, Sumit Madan, Holger Fröhlich

**Affiliations:** Department of Bioinformatics, Fraunhofer Institute for Algorithms and Scientific Computing (SCAI), Schloss Birlinghoven, Sankt Augustin, Germany; Bonn-Aachen International Center for IT (b-it), University of Bonn, Bonn, Germany

## Abstract

Electronic Health Records (EHRs) offer rich data for machine learning, but model generalizability across institutions is hindered by statistical and coding biases. This study investigates domain adaptation (DA) techniques to improve model transfer, focusing on the Ex-Med-BERT transformer architecture for structured EHR data. We compare supervised and unsupervised DA methods in transferring predictive capabilities from the large-scale IBM Explorys database (U.S.) to the UK Biobank. Results across six clinical endpoints show that DA methods outperform fine-tuning, especially with limited target data. These findings emphasize selecting DA strategies based on target data availability and the benefit of incorporating source domain data for robust adaptation.

## 0.1 INTRODUCTION

Machine Learning (ML) models trained to diagnose or prognose diseases have the potential to provide benefit for patients and healthcare providers by aiding clinical decision-making. This may lead to earlier diagnoses, reduced healthcare costs, and improved access to care.[1] Because EHRs provide a comprehensive account of a patient’s medical history that is routinely collected in many healthcare systems, they offer a promising data source for developing ML-based prediction models.

Training prediction models with EHR data faces several challenges. First, differing coding systems and legal frameworks across institutions and countries lead to inconsistencies in disease and procedure documentation, hindering model generalizability across diverse populations.[2], [3] Second, many healthcare settings lack sufficient data for robust model development, particularly for predicting rare diseases.[4] Third, regional variations can result in differences in patient morbidities and medications documented across different EHR systems.

Transfer learning, which fine-tunes a model pre-trained on a related task to predict a new endpoint, is a common approach to address these challenges. When the prediction task is consistent but data distributions differ systematically across domains (e.g., hospitals or countries), domain adaptation (DA) techniques shift the model from the source to the target domain. Originally developed for image analysis, DA is now widely used in fields like natural language processing. Domain Adaptation (DA) addresses distributional shifts by learning domain-invariant representations, where samples with the same label cluster together regardless of their origin.[5] Supervised Domain Adaptation (SDA) utilizes labeled data from both source and target domains, while Unsupervised Domain Adaptation (UDA) relies solely on labeled source data. DA is particularly valuable for EHR data, which can be expected to exhibit substantial domain variability.

Recent studies have explored SDA on a fully connected neural network using MIMIC-III and eCritical ICU datasets.[6] Moreover, DA has been employed to address the temporal domain shift within the MIMIC-IV ICU dataset.[7] While these works highlight the increasing interest in DA for structured EHR data, they are limited in the diversity of DA algorithms considered and do not incorporate recent transformer models for structured EHR data.[8]

This paper explores the application of various unsupervised and semi-supervised domain adaptation (DA) techniques to a foundation model for disease prediction, aiming to overcome existing limitations. We utilize Ex-Med-BERT[9] as our foundation model, which is a BERT-based architecture[10] adapted for structured EHR data. Ex-Med-BERT incorporates diagnoses, medications, and temporal order of doctor visits (encoded as patient age) as input. It also accounts for demographic and regional information and can integrate quantitative lab results when available. Ex-Med-BERT was pre-trained on approximately 3.5 million US patients from the IBM Explorys Therapeutic dataset,[11] encompassing nearly 1 billion records.

This study used Ex-Med-BERT to predict six clinical endpoints across neurological (Alzheimer’s disease, dementia, depression, epilepsy, and Parkinson’s disease), cardiovascular (atrial fibrillation/flutter), and respiratory (chronic obstructive pulmonary disease) conditions. We evaluated three SDA and two UDA strategies to adapt the pre-trained Ex-Med-BERT model to the significantly smaller UK Biobank[12] data. The SDA methods included Classification and Contrastive Semantic Alignment Loss (CCSA),[13] which uses a Siamese network to align semantically similar samples via semantic alignment and separation losses; a triplet loss (TL) approach[14] employing deep metric learning; and Weighted Empirical Risk Minimization (wERM),[15] which uses a weighted loss to optimize source and target data combination for improved generalization. The UDA methods were Minimum Class Confusion (MCC),[16] minimizing target domain prediction ambiguity, and Margin Disparity Discrepancy (MDD),[17] an adversarial DA approach.

## 0.2 METHODS

In this study we compare the performance of five DA approaches to a baseline Ex-Med-BERT model, which was directly applied to the target UK Biobank data after fine-tuning on the source IBM Explorys Therapeutic data.

Notation-wise, we will call the target data *X*_*t*_, where *x*_*t*_ is a vector of the features of one target sample. Similarly, *X*_*s*_ and *x*_*s*_ represent the source data and one source sample, respectively. The model consists of a feature embedding *g* : *X → Z* which maps the input data to a hidden representation, calculated by the transformer layers and the LSTM head, and a classifier *h* : *Z → Y*, which is the classification head that predicts the label from the hidden representation. The complete model is a concatenation of these two functions *f* = *h ° g*.

### 0.2.1 Supervised Domain Adaptation Methods

In SDA, the labels for the target data are available to the model. We are given the source dataset consisting of pairs 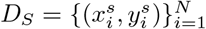 as well as the target dataset consisting of also pairs 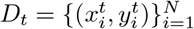. We compare three different SDA approaches, wERM,[15] CCSA[13] and TL.[14]

For the wERM approach, a combined dataset is generated consisting of *n* target samples and *m* source samples 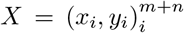. The model is then trained with the original architecture using a weighted loss function where the weights depend on the domain of each sample, as described in Equation (1).

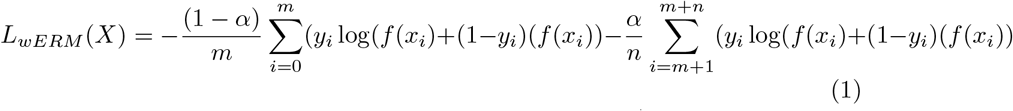

Wang et al. mathematically analyzed the aim of DA to learn invariant representations using heterogeneous data.[15] They argued that in order to adapt the model to the target data and generalize well to the target domain, it is necessary to use a small number of target samples together with the source dataset. The wERM loss function accomplishes this by incorporating both domains in the optimization process, where *α* is treated as a hyperparameter.

The next SDA approach to be discussed is CCSA. In this method, the model learns to pull semantically similar samples closer while pushing dissimilar samples apart by applying two loss functions to the internal representations of both source and target data. These two loss functions, computed on the inner representations of the source and target datasets, are combined with a classification loss calculated on the source data. Together, they form the overall loss that is optimized during training. This approach is implemented in a Siamese network as shown in Figure 1, meaning that the feature embedding *g* is the same for the source and the target dataset, so *g*_*s*_ = *g*_*t*_ = *g*.

**Figure 1:**
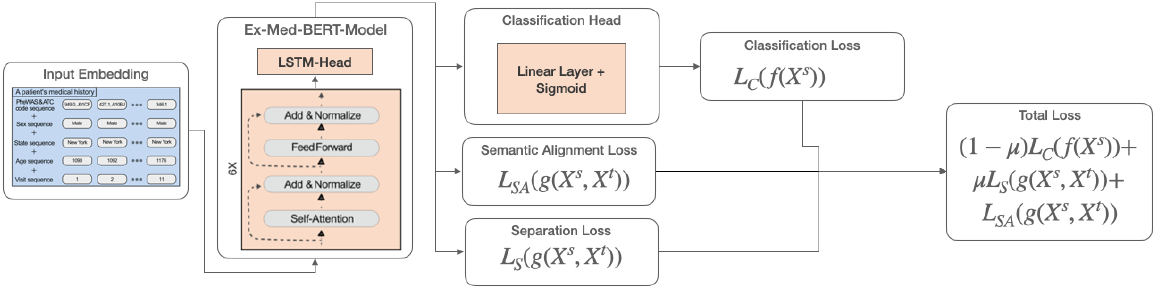
CCSA. Architecture of the CCSA method implemented using a Siamese network. The model combines classification, semantic alignment, and separation losses during optimization.

The semantic alignment loss, defined in Equation (2), ensures that samples sharing the same label across different domains are mapped close together in the feature space Z. The distance measure *d* is provided in (3), where | *·* | denotes the Frobenius norm. *p* (*g*(*X*_*t*_)) and *p* (*g*(*X*_*s*_)) are the probability distributions of the embeddings for the target and source data, respectively.

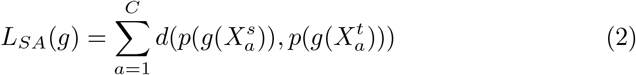

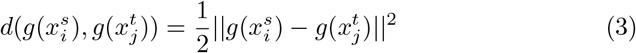

The separation loss, presented in Equation 4, encourages class separation. The model is penalized when the distributions of the embeddings from *X*^*t*^ and *X*^*s*^ for different classes become too similar. Here, *k* is a similarity measure defined in (5). In this measure, *m* is a margin treated as a hyperparameter during training.

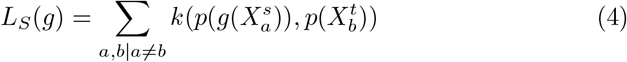

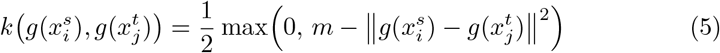

The optimization process combines these representation loss functions with a classification loss computed from the model’s predictions on the source data, as shown in Equation (6), forming the loss function used during training. The factor *µ* is treated as a hyperparameter during this process.

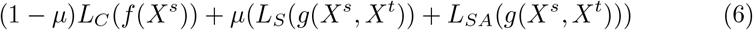

The last SDA approach is TL, which is based on deep metric learning. It aims to train the model to produce embeddings that pull together samples with the same label from different domains while pushing apart samples with different labels. In this approach, the model is trained to generate embeddings that bring together samples with the same label across different domains while pushing apart those with different labels. This promotes domain-invariant feature representations that preserve class-specific structure. To enforce this, the training process constructs all valid triplets from each batch. Each triplet consists of an anchor 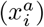, a positive sample 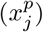 from a different domain but with the same class label, and a negative sample 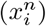 from the same domain as the anchor but with a different label. Formally, the set of valid triplets is defined as 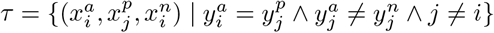, where *i* and *j* denote different domains.

On each of these triplets, the loss function shown in Equation (7) is calculated:

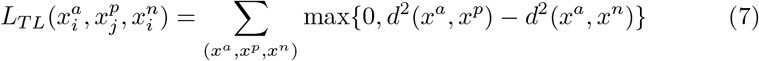

Here, *d* is the Euclidean distance between two embeddings. This function is minimized when the distance between samples from different domains with the same label is less than or equal to the distance between samples from the same domain with different labels. *L*_*TL*_ is calculated for all triplets in *τ*, and its average is used as the loss function during training.

### 0.2.2 Unsupervised Domain Adaptation Methods

While SDA approaches use the class labels of the target dataset, in UDA, the model does not have access to these labels. The source data consists of labelled pairs 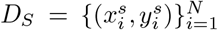, while the target data consists of only unlabelled features 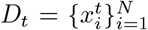. In this study, we compare two UDA approaches: the MCC,[16] and the MDD.[17]

With the MCC approach, the MCC Loss is calculated on the classification of the target dataset as shown in Equation (8):

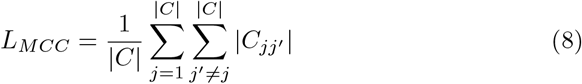

Here, 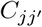 is an estimation of the class confusion between each pair of classes, as detailed in.[16] The function is minimized when no samples are ambiguously classified into two classes simultaneously. The model is trained by minimizing the sum of two terms: (1) the classification loss computed on the source data and (2) the MCC loss, weighted by the hyperparameter *µ*, as shown in Equation 9.

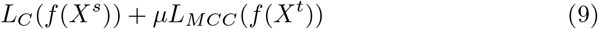

The second SDA approach to be discussed is the MDD, which is an adversarial domain adaptation approach as shown in Figure 2. The model consists of two classification models: the classifier *f*_*c*_, which is the same as in the foundation model and predicts the label of the internal representation of a sample, and the adversary *f*_*a*_, which is an exact copy of the classifier. Before being processed in the adversary, the internal representation is processed in a Gradient Reversal Layer (GRL) which is just the identity in the forward pass but multiplies the gradient with a negative constant in the backward pass. This gradient reversal mechanism encourages the feature distributions of both domains to become as similar as possible, making them indistinguishable to the domain classifier and promoting the learning of domain-invariant features.[18]

**Figure 2:**
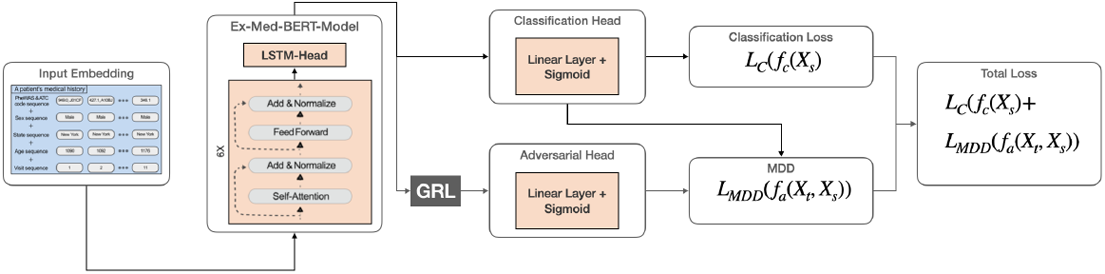
MDD. Structure of the MDD approach using a gradient reversal layer and adversarial classifiers.

The model aims to minimize the maximal difference in expected margin loss of *f*_*a*_ with respect to *f*_*c*_, which is a measure of the discrepancy between the distributions of the embeddings of the source and the target data. If this is minimized, the embeddings should be as domain invariant as possible. To approximate this discrepancy, Equations 10 and 11 are used. While (10) is the standard cross-entropy loss between the predictions of *f*_*a*_ and *f*_*c*_ on *X*_*s*_, (11) is a modified cross-entropy loss between the predictions of the two classification heads on *X*_*t*_. Here, *σ* is the sigmoid function.

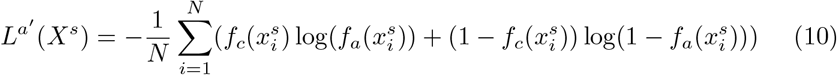

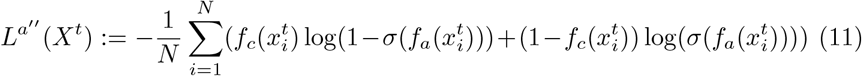

These two equations are combined as shown in Equation (12) to approximate the MDD with the factor *µ* which treated as a hyperparameter during training. Equation (12) is added to the classification loss from *f*_*c*_ on *X*_*s*_, and this sum is minimized during optimization.

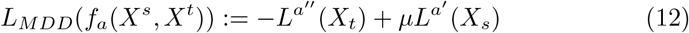

### 0.2.3 Data

- **IBM Explorys Therapeutic (source)** comprises EHR and insurance claims from 4.5 million patients across the USA from 2010 until 2021. For this study, demographic data, drugs, and diagnoses are used.
- **UKBiobank Primary Care Data (target 1)** consists of records from 230,000 patients from the UK. In this study, diagnoses and prescribed drugs are utilized.
- **UKBiobank Hospital Inpatient Data (target 2)** consists of records from 230,000 patients from the UK. In this study, diagnoses and prescribed drugs are utilized.

### 0.2.4 Data Preprocessing

Ex-Med-BERT employs ICD-10 coding for diagnoses and ATC coding for prescribed medications. While clinical events in both the IBM Explorys and UK Biobank Hospital Inpatient datasets are already encoded with these classification systems, the UK Biobank Primary Care dataset uses different coding schemes, Read 2 and CTV3 for diagnoses, and Read 2 for medications. These codes were mapped to ICD-10 and ATC using a mapping table provided by UK Biobank (Resource 592 [19]).

To address the class imbalance present in our data, we implemented propensity score matching (PSM) through the PsmPy library.[20] This approach generates balanced datasets by calculating propensity scores based on patients’ birth year and sex, applying a 1-1 matching technique. In consequence, an adjustment between positive and negative classes with respect to these features for all clinical endpoints was achieved. It is worth to mention that PSM is usually applied in the context of causal inference from observational data. However, here we only aimed for constructing a balanced dataset to avoid potential biases in our subsequent analysis of DA algorithms. Hence, our goal clearly differs from the conventional way in which PSM is applied.

We defined a three-year observation window for each patient. This window spanned the interval [*t*_0_, *t*_1_), where *t*_1_ represented either the date of the patient’s first diagnosis for the endpoint of interest for treatment patients or their last recorded entry in the dataset for the control group. The starting point, *t*_0_, was established as January 1*st* of the year three years prior to *t*_1_. All observations falling within this defined timeframe were included in our analysis.

Table 1 shows the number of patients in the positive class in each dataset. The datasets that are the results of the preprocessing steps explained above are due to the exact balancing of positive and negative classes altogether twice as large.

**Table 1:** Number of patients in the positive class for each clinical endpoint in the IBM Explorys, UK Biobank Primary Care (UKB-GP), and UK Biobank Hospital Inpatient (UKB-INP) datasets.

| Endpoint | IBM Explorys | UKB-GP | UKB-INP |
| --- | --- | --- | --- |
| Alzheimer’s Disease and Dementia | 119,032 | 802 | 3,359 |
| Atrial Fibrillation and Atrial Flutter | 236,191 | 5,922 | 11,860 |
| Chronic Obstructive Pulmonary Disease | 349,992 | 3,958 | 3,240 |
| Depression | 474,700 | 8,102 | 8,905 |
| Epilepsy | 78,957 | 701 | 1,938 |
| Parkinson’s Disease | 151,166 | 290 | 3,100 |

## 0.3 EXPERIMENTAL SETUP

### 0.3.1 Fine-tuning of the baseline model

As mentioned before we used a pre-trained Ex-Med-BERT model, with the pre-training procedure detailed in.[9] The pre-trained transformer layers were then fine-tuned on the source data together with an LSTM-classification head for all clinical endpoints shown in 1.

To optimize the hyperparameters and assessed the model’s performance, a 5-fold nested cross-validation approach is used. The hyperparameters were tuned in the inner cross-validation loop using Bayesian optimization via Optuna.[21]

### 0.3.2 Fine-tuning of the model using Domain Adaptation approaches

We implemented each of the five DA approaches into Ex-Med-BERT. Each model was initialized with parameters previously fine-tuned on the source dataset for its respective endpoint. We trained all DA methods using the same data splits as our baseline model and evaluated performance exclusively on the target test set.

For comparison reasons, we also fine-tuned the pre-trained Ex-Med-BERT foundation model only on target data.

To investigate the impact of target data size, we further tested all DA approaches with a reduced target dataset using only 20% of the available target data during cross-validation while maintaining the same test set as in the full-data experiments.

## 0.4 RESULTS

Figure 3 illustrates endpoint performance on the target data, with detailed results, including confidence intervals, in Appendix A. Performance improvements over the baseline are observed for nearly all endpoints and DA methods. Notably, larger datasets, such as COPD, demonstrate greater improvement compared to smaller datasets like PD.

**Figure 3:**
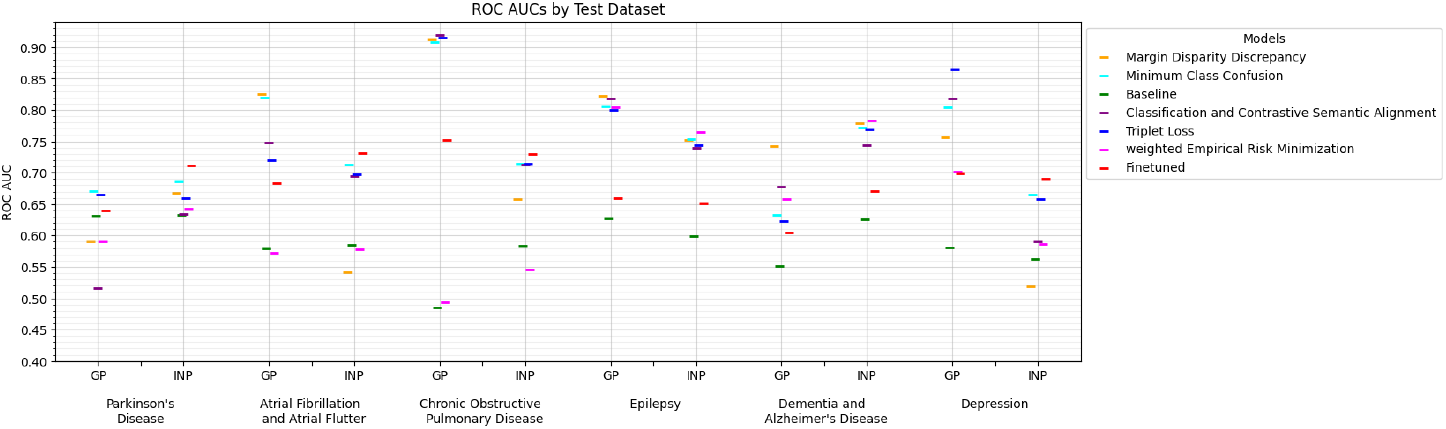
AUROC Scores for full-target setting. AUROC scores across clinical endpoints for each domain adaptation method using the full target dataset. Complex DA techniques outperform simple fine-tuning and wERM in most settings.

Table 2 ranks the improvements by average AUROC increase compared to the baseline. wERM yielded the lowest average increase, while fine-tuning on the target dataset alone doubled that improvement. The domain adaptation methods—MCC, TL, MDD, and CCSA—produced the highest average increases.

**Table 2:**
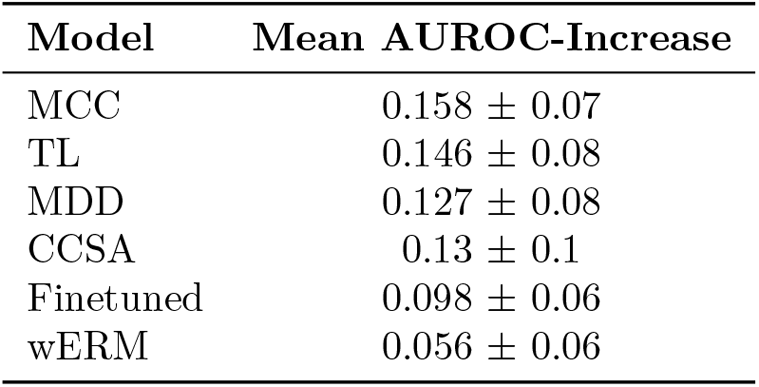
Ranking of the DA-Approaches. Mean AUROC improvements over baseline models for each domain adaptation method, averaged across all endpoints using the full target dataset.

| Model | Mean AUROC-Increase |
| --- | --- |
| MCC | $0.158 \pm 0.07$ |
| TL | $0.146 \pm 0.08$ |
| MDD | $0.127 \pm 0.08$ |
| CCSA | $0.13 \pm 0.1$ |
| Finetuned | $0.098 \pm 0.06$ |
| wERM | $0.056 \pm 0.06$ |

Experiments using 20% of the target data (Figure 4, Table 3), focused on Epilepsy, Dementia and Alzheimer’s Disease, and Depression datasets due to their superior initial performance. While most approaches still improved, gains were smaller compared to using the full target dataset. Simple fine-tuning performed poorly, highlighting the importance of source data for adaptation when target data is limited.

**Table 3:** Ranking of the DA-Approaches. Mean AUROC improvements over baseline models for each domain adaptation method, averaged across all endpoints using the 20% of the target dataset.

| Model | Mean AUROC-Increase |
| --- | --- |
| TL | $0.089 \pm 0.01$ |
| MDD | $0.084 \pm 0.01$ |
| CCSA | $0.06 \pm 0.03$ |
| MCC | $0.054 \pm 0.02$ |
| wERM | $0.05 \pm 0.06$ |
| Finetuned | $0.035 \pm 0.03$ |

**Figure 4:**
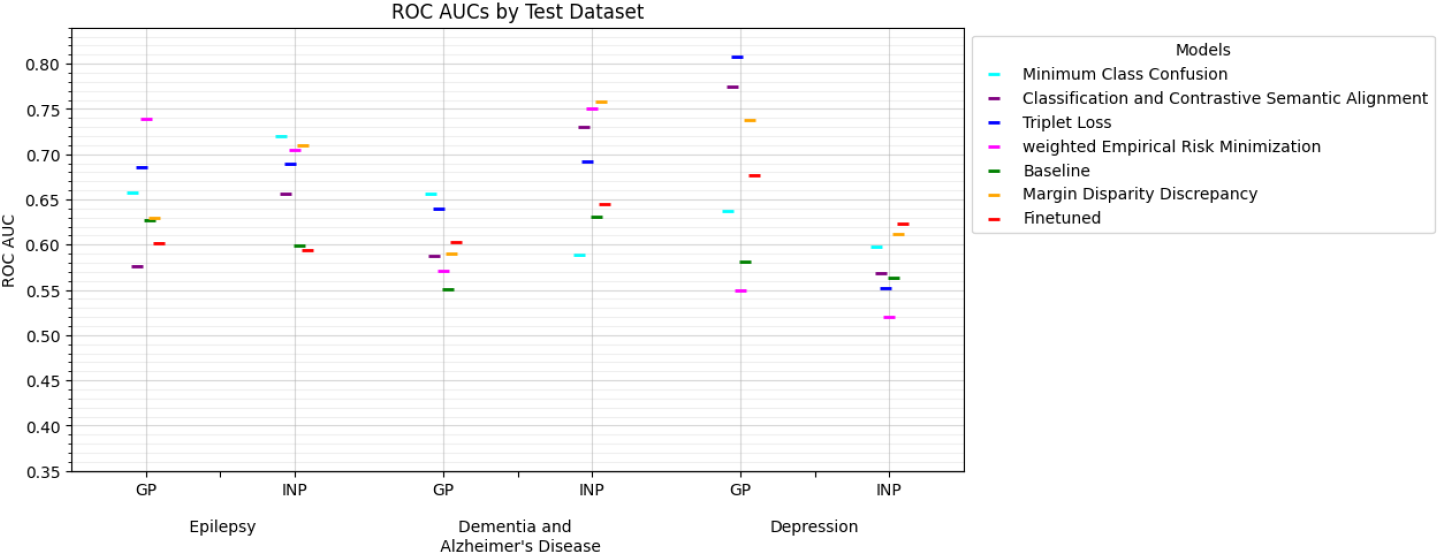
AUROC Scores for the small-target setting. AUROC scores across selected clinical endpoints with only 20% of the target data. Performance gains are reduced compared to full-data settings.

## 0.5 DISCUSSION

Our experiments demonstrate the effectiveness of DA strategies for transferring disease prediction models across EHR datasets. All DA strategies improved performance compared to the baseline, but complex methods that explicitly learn domain-invariant, label-discriminative representations (TL, CCSA, MDD) consistently showed the greatest gains.

TL gains in AUROC were non-proportional to target dataset size. TL improved AUROC by 0.146 with the full dataset, but only 0.089 with 20% of the data – a 60% improvement despite an 80% data reduction. Similar patterns were observed for other DA methods. Notably, wERM achieved consistent gains regardless of target set size (0.061 vs. 0.056). Conversely, fine-tuning on target data alone yielded performance gains proportional to target set size (0.095 for the full set and 0.035 for the reduced set). These findings demonstrate the importance of leveraging source data for effective domain adaptation, particularly when limited target data is available.

These results indicate that the optimal DA method depends on data availability. With large target datasets, complex methods like TL, MCC, CCSA, or MDD provide the greatest performance improvement. Conversely, wERM remains relatively robust with limited target data, albeit with slightly reduced effectiveness.

## 0.6 CONCLUSION

Our study is the first to systematically evaluate various SDA and UDA techniques for enhancing the cross-domain generalization of a transformer-based model using structured EHR data. Experiments across six clinical endpoints demonstrate that domain adaptation strategies promoting domain-invariant embeddings – such as TL, CCSA, MCC, and MDD – surpass both simple finetuning and wERM, especially with ample labeled target data. In low-target data scenarios, incorporating source domain information effectively mitigated performance decline. These results emphasize the crucial relationship between domain adaptation method complexity and target data availability, indicating that practical data constraints should guide method selection.

## Data Availability

All data produced in the present work are contained in the manuscript

## FUNDING AND ACKNOWLEDGEMENTS

This project has received funding from the European Union’s Horizon Europe research and innovation programme under grant agreement No [101112135] (Integration of heterogeneous Data and Evidence towards Regulatory and HTA Acceptance [IDERHA]) through the Innovative Health Initiative (IHI) Joint Undertaking (JU). Support is also received from life science industries represented by COCIR, EFPIA / Vaccines Europe, EuropaBio and MedTech Europe. Support is also received from our Swiss and UK partners.

This research has been conducted using the UK Biobank resource under application 67829.

## A Results Tables

**Table 4:** Evaluation Results. This table displays the outcomes obtained from the INP dataset in the small-target setting, highlighting the optimal values for each endpoint in bold.

| Endpoint | Models | AUROC | AUPR | $\Delta$ AUROC | $\Delta$ PRAUC |
| --- | --- | --- | --- | --- | --- |
| EPI | Baseline | $0.600 \pm 0.000$ | $0.483 \pm 0.000$ | 0.000 | 0.000 |
| | Finetuned | $0.595 \pm 0.027$ | $0.546 \pm 0.027$ | -0.023 | 0.043 |
| | CCSA | $0.657 \pm 0.063$ | $0.555 \pm 0.078$ | 0.039 | 0.052 |
| | TL | $0.690 \pm 0.035$ | $0.583 \pm 0.047$ | 0.073 | 0.080 |
| | wERM | $0.704 \pm 0.039$ | $0.622 \pm 0.051$ | 0.087 | 0.119 |
| | MDD | $0.709 \pm 0.034$ | $0.623 \pm 0.044$ | 0.092 | 0.120 |
|  | MCC | **$0.719 \pm 0.016$** | **$0.641 \pm 0.026$** | 0.102 | 0.138 |
| DEM_AD | Baseline | $0.630 \pm 0.000$ | $0.543 \pm 0.000$ | 0.000 | 0.000 |
| | Finetuned | $0.645 \pm 0.020$ | $0.611 \pm 0.014$ | 0.015 | 0.067 |
| | CCSA | $0.730 \pm 0.020$ | $0.675 \pm 0.021$ | 0.100 | 0.132 |
| | TL | $0.692 \pm 0.027$ | $0.624 \pm 0.035$ | 0.062 | 0.081 |
| | wERM | $0.751 \pm 0.009$ | $0.702 \pm 0.009$ | 0.120 | 0.158 |
|  | MDD | **$0.758 \pm 0.021$** | **$0.716 \pm 0.028$** | 0.128 | 0.172 |
| | MCC | $0.589 \pm 0.087$ | $0.526 \pm 0.090$ | -0.041 | -0.017 |
| DE | Baseline | $0.563 \pm 0.000$ | $0.496 \pm 0.000$ | 0.000 | 0.000 |
|  | Finetuned | **$0.623 \pm 0.028$** | **$0.567 \pm 0.021$** | 0.061 | 0.071 |
| | CCSA | $0.568 \pm 0.015$ | $0.494 \pm 0.021$ | 0.005 | -0.002 |
| | TL | $0.552 \pm 0.008$ | $0.476 \pm 0.011$ | -0.011 | -0.021 |
| | wERM | $0.520 \pm 0.042$ | $0.447 \pm 0.043$ | -0.043 | -0.049 |
| | MDD | $0.612 \pm 0.026$ | $0.528 \pm 0.023$ | 0.049 | 0.031 |
| | MCC | $0.597 \pm 0.012$ | $0.541 \pm 0.017$ | 0.035 | 0.044 |

**Table 5:** Evaluation Results. This table displays the outcomes obtained from the GP dataset in the small-target setting, highlighting the optimal values for each endpoint in bold.

| Endpoint | Models | AUROC | AUPR | $\Delta$ AUROC | $\Delta$ PRAUC |
| --- | --- | --- | --- | --- | --- |
| EPI | Baseline | $0.627 \pm 0.000$ | $0.448 \pm 0.000$ | 0.000 | 0.000 |
| | Finetuned | $0.602 \pm 0.012$ | $0.389 \pm 0.026$ | 0.012 | 0.031 |
| | CCSA | $0.576 \pm 0.098$ | $0.414 \pm 0.101$ | -0.014 | 0.056 |
| | TL | $0.685 \pm 0.050$ | $0.534 \pm 0.105$ | 0.096 | 0.176 |
|  | wERM | **$0.739 \pm 0.026$** | **$0.648 \pm 0.034$** | 0.149 | 0.290 |
| | MDD | $0.630 \pm 0.068$ | $0.448 \pm 0.113$ | 0.040 | 0.090 |
| | MCC | $0.658 \pm 0.101$ | $0.534 \pm 0.162$ | 0.068 | 0.176 |
| DEM_AD | Baseline | $0.551 \pm 0.000$ | $0.529 \pm 0.000$ | 0.000 | 0.000 |
| | Finetuned | $0.603 \pm 0.024$ | $0.578 \pm 0.032$ | 0.054 | 0.079 |
| | CCSA | $0.588 \pm 0.034$ | $0.566 \pm 0.034$ | 0.039 | 0.067 |
| | TL | $0.639 \pm 0.056$ | $0.622 \pm 0.035$ | 0.090 | 0.124 |
| | wERM | $0.570 \pm 0.024$ | $0.550 \pm 0.036$ | 0.021 | 0.052 |
| | MDD | $0.590 \pm 0.031$ | $0.568 \pm 0.027$ | 0.041 | 0.070 |
|  | MCC | **$0.656 \pm 0.022$** | **$0.649 \pm 0.040$** | 0.107 | 0.150 |
| DE | Baseline | $0.581 \pm 0.000$ | $0.475 \pm 0.000$ | 0.000 | 0.000 |
| | Finetuned | $0.676 \pm 0.015$ | $0.569 \pm 0.012$ | 0.095 | 0.095 |
| | CCSA | $0.774 \pm 0.036$ | $0.689 \pm 0.051$ | 0.193 | 0.215 |
|  | TL | **$0.808 \pm 0.004$** | **$0.732 \pm 0.011$** | 0.227 | 0.258 |
| | wERM | $0.549 \pm 0.108$ | $0.448 \pm 0.103$ | -0.032 | -0.027 |
| | MDD | $0.738 \pm 0.021$ | $0.655 \pm 0.030$ | 0.157 | 0.180 |
| | MCC | $0.638 \pm 0.094$ | $0.620 \pm 0.105$ | 0.057 | 0.145 |

**Table 6:** Evaluation Results. This table displays the outcomes obtained from the INP dataset in the full-target setting, highlighting the optimal values for each endpoint in bold.

| Endpoint | Models | AUROC | AUPR | $\Delta$ AUROC | $\Delta$ PRAUC |
| --- | --- | --- | --- | --- | --- |
| PD | Baseline | $0.632 \pm 0.000$ | $0.460 \pm 0.000$ | 0.000 | 0.000 |
|  | Finetuned | **$0.711 \pm 0.020$** | **$0.571 \pm 0.016$** | 0.079 | 0.111 |
| | CCSA | $0.634 \pm 0.140$ | $0.510 \pm 0.126$ | 0.002 | 0.050 |
| | TL | $0.66 \pm 0.041$ | $0.482 \pm 0.054$ | 0.028 | 0.022 |
| | wERM | $0.643 \pm 0.019$ | $0.455 \pm 0.028$ | 0.011 | -0.005 |
| | MDD | $0.668 \pm 0.050$ | $0.509 \pm 0.083$ | 0.035 | 0.049 |
| | MCC | $0.686 \pm 0.043$ | $0.547 \pm 0.033$ | 0.053 | 0.087 |
| AFF | Baseline | $0.586 \pm 0.000$ | $0.455 \pm 0.000$ | 0.000 | 0.000 |
|  | Finetuned | **$0.731 \pm 0.003$** | **$0.649 \pm 0.004$** | 0.145 | 0.194 |
| | CCSA | $0.695 \pm 0.014$ | $0.604 \pm 0.021$ | 0.109 | 0.149 |
| | TL | $0.697 \pm 0.009$ | $0.577 \pm 0.024$ | 0.111 | 0.121 |
| | wERM | $0.578 \pm 0.008$ | $0.452 \pm 0.007$ | -0.007 | -0.003 |
| | MDD | $0.542 \pm 0.143$ | $0.464 \pm 0.120$ | -0.044 | 0.009 |
| | MCC | $0.712 \pm 0.009$ | $0.627 \pm 0.008$ | 0.127 | 0.171 |
| COPD | Baseline | $0.584 \pm 0.000$ | $0.482 \pm 0.000$ | 0.000 | 0.000 |
|  | Finetuned | **$0.730 \pm 0.014$** | **$0.650 \pm 0.014$** | 0.146 | 0.167 |
| | CCSA | $0.713 \pm 0.014$ | $0.636 \pm 0.011$ | 0.129 | 0.153 |
| | TL | $0.714 \pm 0.007$ | $0.627 \pm 0.014$ | 0.130 | 0.145 |
| | wERM | $0.545 \pm 0.021$ | $0.454 \pm 0.017$ | -0.039 | -0.028 |
| | MDD | $0.658 \pm 0.077$ | $0.573 \pm 0.064$ | 0.074 | 0.091 |
| | MCC | $0.714 \pm 0.020$ | $0.630 \pm 0.021$ | 0.130 | 0.148 |
| EPI | Baseline | $0.600 \pm 0.000$ | $0.483 \pm 0.000$ | 0.000 | 0.000 |
| | Finetuned | $0.651 \pm 0.025$ | $0.607 \pm 0.025$ | 0.051 | 0.123 |
| | CCSA | $0.739 \pm 0.033$ | $0.656 \pm 0.051$ | 0.139 | 0.173 |
| | TL | $0.744 \pm 0.021$ | $0.667 \pm 0.027$ | 0.144 | 0.184 |
|  | wERM | **$0.765 \pm 0.015$** | **$0.693 \pm 0.014$** | 0.165 | 0.209 |
| | MDD | $0.752 \pm 0.021$ | $0.682 \pm 0.021$ | 0.152 | 0.198 |
| | MCC | $0.753 \pm 0.027$ | $0.673 \pm 0.039$ | 0.153 | 0.190 |
| DEM AD | Baseline | $0.626 \pm 0.000$ | $0.541 \pm 0.000$ | 0.000 | 0.000 |
| | Finetuned | $0.671 \pm 0.015$ | $0.640 \pm 0.019$ | 0.044 | 0.099 |
| | CCSA | $0.743 \pm 0.030$ | $0.690 \pm 0.038$ | 0.117 | 0.149 |
| | TL | $0.769 \pm 0.020$ | $0.705 \pm 0.022$ | 0.143 | 0.165 |
|  | wERM | **$0.783 \pm 0.019$** | **$0.743 \pm 0.018$** | 0.157 | 0.202 |
| | MDD | $0.778 \pm 0.010$ | $0.739 \pm 0.015$ | 0.152 | 0.198 |
| | MCC | $0.771 \pm 0.019$ | $0.737 \pm 0.014$ | 0.145 | 0.196 |
| DE | Baseline | $0.563 \pm 0.000$ | $0.496 \pm 0.000$ | 0.000 | 0.000 |
| | Finetuned | $0.691 \pm 0.011$ | $0.631 \pm 0.008$ | 0.128 | 0.134 |
| | CCSA | $0.591 \pm 0.040$ | $0.526 \pm 0.025$ | 0.028 | 0.029 |
| | TL | $0.659 \pm 0.016$ | $0.575 \pm 0.013$ | 0.096 | 0.079 |
| | wERM | $0.587 \pm 0.093$ | $0.533 \pm 0.082$ | 0.024 | 0.036 |
| | MDD | $0.519 \pm 0.083$ | $0.448 \pm 0.067$ | -0.044 | -0.049 |
|  | MCC | **$0.665 \pm 0.012$** | **$0.605 \pm 0.013$** | 0.102 | 0.109 |

**Table 7:** Evaluation Results. This table displays the outcomes obtained from the GP dataset in the full-target setting, highlighting the optimal values for each endpoint in bold.

| Endpoint | Models | AUROC | AUPR | $\Delta$ AUROC | $\Delta$ PRAUC |
| --- | --- | --- | --- | --- | --- |
| PD | Baseline | $0.632 \pm 0.000$ | $0.609 \pm 0.000$ | 0.000 | 0.000 |
| | Finetuned | $0.639 \pm 0.040$ | $0.623 \pm 0.069$ | 0.007 | 0.014 |
| | CCSA | $0.516 \pm 0.079$ | $0.508 \pm 0.073$ | -0.116 | -0.101 |
| | TL | $0.665 \pm 0.046$ | $0.645 \pm 0.048$ | 0.034 | 0.036 |
| | wERM | $0.590 \pm 0.027$ | $0.567 \pm 0.043$ | -0.041 | -0.043 |
| | MDD | $0.591 \pm 0.138$ | $0.579 \pm 0.115$ | -0.041 | -0.031 |
|  | MCC | **$0.671 \pm 0.034$** | **$0.653 \pm 0.047$** | 0.039 | 0.044 |
| AFF | Baseline | $0.580 \pm 0.000$ | $0.562 \pm 0.000$ | 0.000 | 0.000 |
| | Finetuned | $0.684 \pm 0.016$ | $0.669 \pm 0.011$ | 0.104 | 0.107 |
| | CCSA | $0.748 \pm 0.104$ | $0.731 \pm 0.112$ | 0.168 | 0.169 |
| | TL | $0.721 \pm 0.111$ | $0.691 \pm 0.104$ | 0.141 | 0.130 |
| | wERM | $0.572 \pm 0.009$ | $0.552 \pm 0.008$ | -0.008 | -0.010 |
|  | MDD | **$0.825 \pm 0.034$** | **$0.817 \pm 0.035$** | 0.245 | 0.256 |
| | MCC | $0.819 \pm 0.011$ | $0.812 \pm 0.010$ | 0.239 | 0.250 |
| COPD | Baseline | $0.485 \pm 0.000$ | $0.511 \pm 0.000$ | 0.000 | 0.000 |
| | Finetuned | $0.753 \pm 0.016$ | $0.759 \pm 0.022$ | 0.267 | 0.248 |
|  | CCSA | **$0.919 \pm 0.005$** | **$0.922 \pm 0.008$** | 0.434 | 0.411 |
| | TL | $0.915 \pm 0.015$ | $0.899 \pm 0.036$ | 0.430 | 0.388 |
| | wERM | $0.494 \pm 0.015$ | $0.520 \pm 0.014$ | 0.009 | 0.009 |
| | MDD | $0.912 \pm 0.006$ | $0.920 \pm 0.005$ | 0.427 | 0.409 |
| | MCC | $0.907 \pm 0.015$ | $0.918 \pm 0.012$ | 0.422 | 0.407 |
| EPI | Baseline | $0.627 \pm 0.000$ | $0.448 \pm 0.000$ | 0.000 | 0.000 |
| | Finetuned | $0.659 \pm 0.018$ | $0.491 \pm 0.019$ | 0.032 | 0.043 |
| | CCSA | **$0.818 \pm 0.019$** | $0.740 \pm 0.025$ | 0.191 | 0.292 |
| | TL | $0.799 \pm 0.034$ | $0.711 \pm 0.034$ | 0.172 | 0.263 |
| | wERM | $0.805 \pm 0.032$ | **$0.732 \pm 0.025$** | 0.178 | 0.284 |
| | MDD | $0.823 \pm 0.022$ | $0.742 \pm 0.029$ | 0.196 | 0.294 |
| | MCC | $0.806 \pm 0.020$ | $0.715 \pm 0.034$ | 0.179 | 0.268 |
| DEM_AD | Baseline | $0.551 \pm 0.000$ | $0.529 \pm 0.000$ | 0.000 | 0.000 |
| | Finetuned | $0.604 \pm 0.013$ | $0.573 \pm 0.022$ | 0.053 | 0.044 |
| | CCSA | $0.678 \pm 0.056$ | $0.654 \pm 0.066$ | 0.128 | 0.124 |
| | TL | $0.623 \pm 0.168$ | $0.621 \pm 0.134$ | 0.072 | 0.092 |
| | wERM | $0.658 \pm 0.162$ | $0.688 \pm 0.156$ | 0.107 | 0.158 |
|  | MDD | **$0.742 \pm 0.021$** | **$0.751 \pm 0.022$** | 0.192 | 0.222 |
| | MCC | $0.632 \pm 0.142$ | $0.669 \pm 0.126$ | 0.082 | 0.140 |
| DE | Baseline | $0.581 \pm 0.000$ | $0.475 \pm 0.000$ | 0.000 | 0.000 |
| | Finetuned | $0.699 \pm 0.008$ | $0.582 \pm 0.012$ | 0.118 | 0.107 |
| | CCSA | $0.818 \pm 0.083$ | $0.781 \pm 0.063$ | 0.237 | 0.307 |
|  | TL | **$0.865 \pm 0.009$** | **$0.824 \pm 0.020$** | 0.284 | 0.350 |
| | wERM | $0.701 \pm 0.275$ | $0.700 \pm 0.227$ | 0.120 | 0.225 |
| | MDD | $0.757 \pm 0.050$ | $0.666 \pm 0.075$ | 0.176 | 0.191 |
| | MCC | $0.804 \pm 0.050$ | $0.806 \pm 0.034$ | 0.223 | 0.332 |

